# Comparing Client and Provider Preferences for HIV Care Coordination Program Features using Discrete Choice Experiments

**DOI:** 10.1101/2024.09.15.24313701

**Authors:** Rebecca Zimba, Chunki Fong, Madellena Conte, Honoria Guarino, Tigran Avoundjian, Jennifer Carmona, Grace Herndon, Gina Gambone, Mary Irvine, Denis Nash

## Abstract

**Introduction:** The New York City (NYC) HIV Care Coordination Program (CCP) is designed to promote care engagement and treatment adherence among the people with HIV (PWH) who struggle the most with these key components of managing their health. We assessed preferences for CCP components among PWH enrolled in the program (“clients”) and among providers of CCP services. In this report we compare the preferences between clients and providers, previously analyzed separately.

**Methods:** We used a discrete choice experiment to assess preferences for four CCP features (“attributes”): Help with Adherence to Antiretroviral Therapy (ART), Help with Primary Care Appointments, Help with Issues other than Primary Care, and Where Program Visits Happen. Each of these attributes had three to four variants (“levels”). In the original surveys, levels within Where Program Visits Happen varied by participant type (client versus provider). We re-coded the levels by visit location (VL) or by travel time (TT) to make them comparable, and report results from both approaches. Preferences were quantified using the relative importance of the attributes and utility of the levels.

**Results:** January 2020 to March 2021, 152 providers and 181 clients completed the survey. In both the VL and TT analyses, clients were most influenced by Help with Adherence to ART, preferring medication reminders via phone or text, and Where Program Visits Happen, preferring visits via phone or video chat. In the VL analysis, providers were most influenced by Help with Adherence to ART, valuing directly observed therapy most highly, and Help with Issues other than Primary Care, valuing helping clients with connections to specialty medical care. In the TT analysis, providers were most influenced by Where Program Visits Happen, preferring to meet at clients’ homes, and Help with Issues other than Primary Care, again preferring helping clients make connections to specialty medical care.

**Conclusions:** Client and provider preferences clearly diverged with regard to CCP service intensity: in the aggregate, clients tended to prefer lower-intensity services, whereas providers endorsed higher-intensity services. These results highlight the importance of engaging clients as partners in decisions about program services so that they are aligned with client values.

## Introduction

Ryan White Part A-funded (RWPA) HIV Care Coordination Programs (CCPs) in New York City (NYC) aim to help the most vulnerable persons with HIV (PWH) to consistently engage in care and adhere to their HIV treatment regimens. The CCP incorporates outreach, case management, interdisciplinary case conferences, patient navigation, antiretroviral therapy (ART) adherence support, and structured health promotion. These programs are either co-located with primary medical care or formally affiliated with primary medical care providers. Originally launched in 2009 by the NYC Department of Health and Mental Hygiene (NYC Health Department) [1], the CCP underwent revisions in 2018 to reduce challenges to implementation, improve client engagement, and enhance the program’s delivery and impact.

That same year, the NYC Health Department and City University of New York Institute for Implementation Science in Population Health launched the PROMISE study (<u>P</u>rogram <u>R</u>efinements to <u>O</u>ptimize <u>M</u>odel Impact and <u>S</u>calability Based on <u>E</u>vidence) to evaluate the implementation and impact of revisions to the CCP [2]. As part of the PROMISE study, we conducted discrete choice experiments (DCEs) among CCP clients and service providers to better understand their preferences for different components of the program. In DCEs participants are presented with a series of choices between sets of hypothetical programs, services, or products made up of predefined features that vary within and across choice sets. We have separately reported findings from the client and provider DCEs [3–5].

Other studies have found discordance between preferences of physicians and patients (or patients’ caregivers/guardians) for the treatment of various conditions, including asthma, diabetes, cancer, and psoriasis [6–11]. However, there is a dearth of research comparing preferences of providers and clients of HIV care coordination or medical case management programs for the support and services they provide or receive, respectively. Discordant client-provider preferences may indicate diverging opinions about the value of care coordination components, which could impact client satisfaction, provider-client communication, and treatment adherence [12,13]. Here we aim to compare CCP clients’ and providers’ preferences as measured through DCEs.

## Methods

### Population and recruitment

The target population and recruitment methods have been described elsewhere [4,14]. Briefly, we conducted a census of NYC RWPA providers in the core CCP roles of patient navigators/health educators, care coordinators/case managers, and program directors or other administrators at the 25 CCP-implementing agencies, and invited all of these providers to participate with the survey URL and a unique identification code. CCP clients were eligible for the client DCE if they were ≥18 years old, spoke English, Spanish, or French as their primary or secondary language, and were enrolled in the CCP at one of six partnering agencies. Clients were recruited by staff of the partnering CCP program in which they were being served. Updated lists of eligible clients were shared with staff during data collection to ensure that clients more newly enrolled in the CCP were included. Staff provided clients with unique identification codes if they agreed to participate. All participants received a $25 gift card upon survey completion. The NYC Health Department IRB approved the study.

### DCE design

Our process for designing and implementing the DCE has been described elsewhere [3,4,14]; a summary is provided here. We conducted focus groups with providers and clients to come up with a list of possible CCP features that could be included in the DCEs for both populations to facilitate comparison. The study team reviewed notes and transcripts from the focus groups internally and with the PROMISE Study Advisory Board. To be good candidates for the DCE, the features needed to be salient to the aims of the CCP and flexible in their focus (i.e., type of service), intensity (i.e., frequency of service or degree of interaction), or mode of delivery (i.e., location or in-person versus virtual). Our final design included four features, called “attributes” in DCE design: Help with Adherence to ART; Help with Primary Care Appointments; Help with Issues other than Primary Care; and Where Program Visits Happen. Each attribute included three to four “levels.” Levels were illustrated with black-and-white graphics to facilitate comprehension and comparison across the choice alternatives. We customized the wording of the levels as needed for the client and provider audiences. See Supplemental Figure 1 and Supplemental Table 1.

We designed the survey using Lighthouse Studio Version 9.8.1 (Sawtooth Software, Provo, UT, USA) and deployed the survey via Sawtooth’s online hosting platform. The DCE included 10 comparison tasks, with two hypothetical program options per task. We used Sawtooth’s Balanced Overlap method [15,16] to generate random tasks in which each level within an attribute appeared about the same number of times as the other levels within that attribute (i.e., level balance), the same level within an attribute could sometimes appear in both hypothetical options in a single task, and levels in different attributes appeared independently of one another (i.e. orthogonality). The survey was deployed in English for providers and in English, Spanish, and French for clients.

In each choice exercise, we prompted providers and clients: “Imagine that you had to choose between two programs with the features below. Select the one that you would prefer.” Though the DCE presents mutually exclusive options in the DCE task format, all levels are supported by the CCP. Relatedly, we did not include a “None” option, since everyone who took the DCE was either a client or provider of the CCP and would necessarily engage with some or all of the features present in each alternative. Additional descriptive and clinical data were collected after the choice exercises or merged from contract documents or other NYC Health Department records, including site-specific variables such as whether the site was co-located in a medical facility.

### Sample size and data collection

To calculate minimum sample sizes, we used n ≥ (500c)/(ta), where n is the number of participants, 500 is the minimum number of times each main-effect level appears in the design, c is the maximum number of levels among all of the attributes, t is the number of choice tasks, and a is the number of options per task [15,16]. Given a maximum of four levels, 10 choice tasks, and two alternatives per task (*c*=4, *t*=10, and *a*=2) the minimum sample size is 100. We aimed to obtain 150 responses from providers and 200 responses from clients.

In early January 2020, we emailed personalized links to 227 eligible providers. The provider survey was closed in early March 2020. The client DCE remained open from late January 2020 until March 2021 due to challenges recruiting participants during the COVID-19 pandemic. A total of 884 clients were eligible for the DCE during the recruitment period.

### Analysis

#### Attributes and levels

The attributes and levels in both DCEs needed to be as similar as possible to facilitate comparison between the provider and the client results. In the original DCEs, the levels were comparable for all attributes except Where Program Visits Happen, since providers travel to the clients’ homes for home visits, while clients travel to the CCP site when meeting at the program. In one analysis, we retained the original focus of the attribute (visit location) and combined the levels with the same location: the clients’ home, by phone or video chat, and the program location. In the other analysis, rather than focusing on visit location, we mapped levels across surveys according to travel time, i.e., 30 or 60 minutes to meet at client’s home (for providers) corresponding to 30 or 60 minutes to meet at the program (for clients). Meeting at the program (for providers) and meeting at home (for clients) mapped to no travel. The level for meeting by phone or video chat required no adjusting.

We have described our analytical approaches elsewhere [3–5]. Briefly, using a hierarchical Bayesian multinomial logit model, we estimated relative importances, which are a measures of attributes’ influence on choice behaviors, and part-worth utilities, which are measures of preference for levels within an attribute; utilities were zero-centered [17,18]. We used the independence t-test to compare group differences for continuous variables, and the chi-square test or Fisher’s Exact test for categorical variables, where applicable.

#### Sensitivity analyses

On March 20, 2020, New York State Governor Andrew Cuomo signed the “New York State on PAUSE” executive order, and many of our partnering agencies transitioned to telehealth at or around that time [19]. As a sensitivity analysis, we restricted the data to responses completed before the suspension of in-person services. Results are reported in the Supplemental File.

#### Software

Importances and utilities were estimated using Sawtooth Software’s Lighthouse Studio 9.8.1. Additional analyses were done using SAS (Release 9.4 SAS Institute Inc., Cary, NC, USA) and R (version 4.3.2).

## Results

Surveys were completed by 181 clients (January 2020 - March 2021) and 152 providers (January - March 2020). Table 1 presents basic demographic characteristics for both groups. Most of the providers (52%) were under 40 years old, while most of the clients (60.2%) were 50 or older. Almost half of the providers identified as Latino/a, and two-third of the clients (66.9%) identified as Black non-Hispanic. Most of the providers and clients identified as women (68.4% and 55.3%, respectively). All 152 providers and 140 (77%) of clients completed the DCE before the suspension of in-person services due to the COVID-19 pandemic. Additional characteristics are summarized in our earlier client- and provider-specific papers [3–5]; notably 92.3% of clients had been enrolled in the CCP for more than 1 year, 78.5% were virally suppressed, 68.5% were stably housed, and 26.5% had received DOT services within the CCP.

**Table 1.** Demographic characteristics of Clients and Providers.

|  | **Clients**<br>(n=181)<br>n (%) | **Providers**<br>(n=152)<br>n (%) |
| --- | --- | --- |
| **Age group (years)** |  |  |
| < 40 | 38 (21.0) | 79 (52.0) |
| 40-49 | 34 (18.8) | 32 (21.1) |
| 50-59 | 55 (30.4) | 29 (19.1) |
| ≥ 60 | 54 (29.8) | 12 (7.9) |
| **Race/ethnicity** |  |  |
| Hispanic | 51 (28.2) | 74 (48.7) |
| Black non-Hispanic | 121 (66.9) | 51 (33.6) |
| White non-Hispanic | 5 (2.8) | 12 (7.9) |
| American Indian/Alaska Native, Asian/Pacific Islander | 4 (2.2) | 6 (3.9) |
| Multi-racial, other, or unreported | 0 (0.0) | 9 (5.9) |
| **Gender identity** |  |  |
| Woman | 100 (55.3) | 104 (68.4) |
| Man | 80 (44.2) | 43 (28.3) |
| Transgender or other identity | 1 (0.6) | 5 (3.4) |

### Importances

Figure 1 compares the average relative importances for each attribute between providers and clients. Panel A shows the relative importances when levels in the Where Program Visits Happen attribute were combined by location. All the attribute importance comparisons between provider and client responses were statistically significant. The most important attribute for clients was Help with Adherence to ART (30.5% [95% CI 28.5%, 32.4%]), followed by Where Program Visits Happen (visit location) (26.8% [24.9%, 28.6%]), Help with Issues other than Primary Care (24.1% [22.4%, 25.7%]), and Help with Primary Care Appointments (18.7% [17.5%, 20%]). The most important attribute for providers was Help with Issues other than Primary Care (26.9% [25.2%, 28.6%]), closely followed by Help with Adherence to ART (25.5% [23.5%, 27.5%]), Help with Primary Care Appointments (25.3% [23.8%, 26.9%]), and Where Program Visits Happen (visit location) (22.3% [20.7%, 23.9%]). The range of importances for clients (18.7% - 30.5%, 11.8 percentage points [pp]) was larger than for the providers (22.3% - 26.9%, 4.6pp). Help With Primary Care Appointments had the biggest difference in relative importances between clients and providers (6.6pp lower among clients [−8.6pp, −4.7pp]).

**Figure 1.**
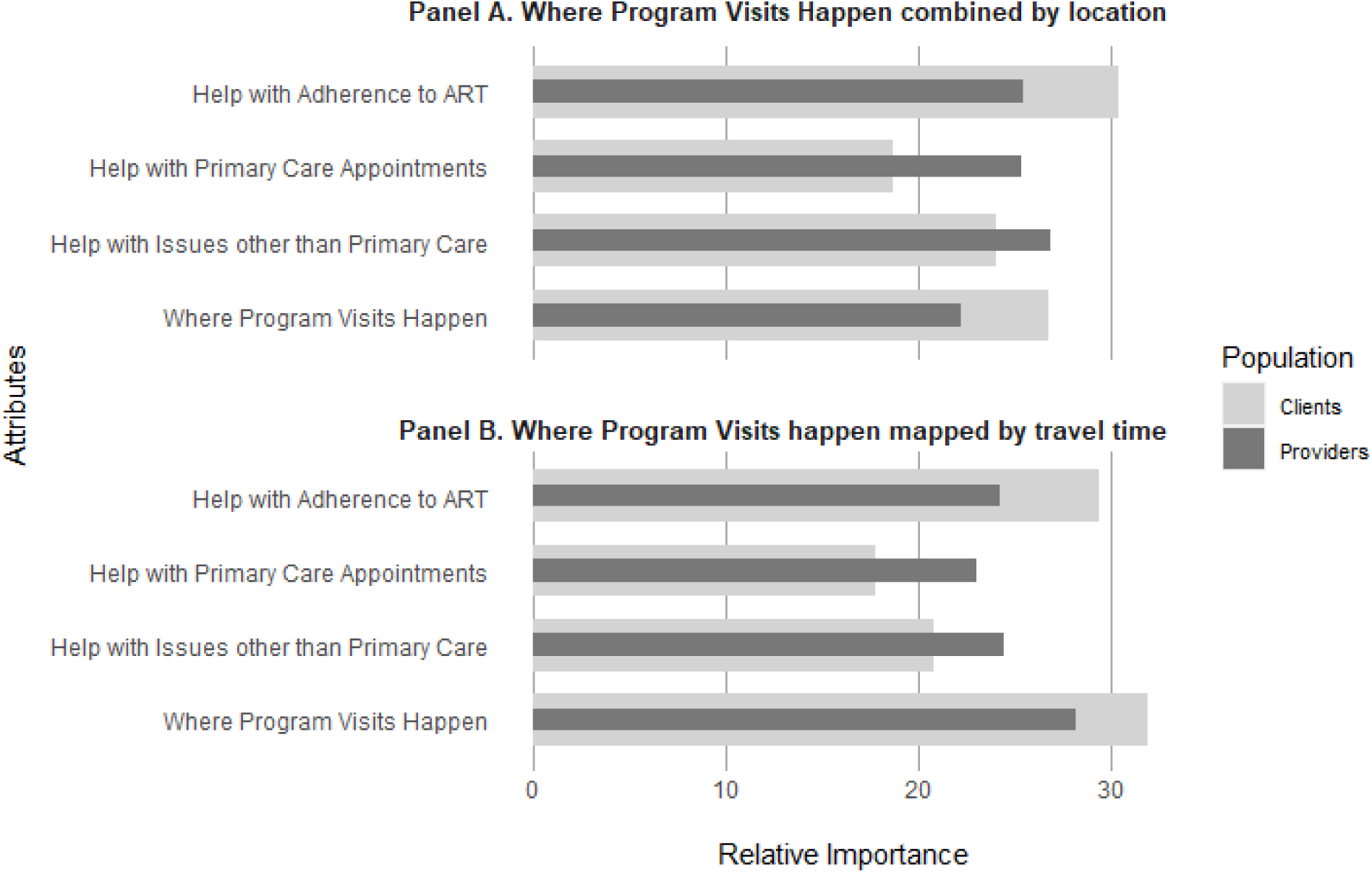
Attribute relative importances by analysis and population.

Figure 1, Panel B compares the average relative importances for each attribute between providers and clients when levels in the Where Program Visits Happen attribute were mapped by travel time. All the attribute importance comparisons between provider and client responses were again statistically significant. Among clients, the most important attribute was Where Program Visits Happen (travel time) (32.0% [30.1%, 33.8%]), followed by Help with Adherence to ART (29.4% [27.5%, 31.4%]), Help with Issues other than Primary Care (20.8% [19.5%, 22.2%]), and Help with Primary Care Appointments (17.8% [16.6%, 19.0%]). Among providers, Where Program Visits Happen (travel time) was also the most important attribute (28.2% [26.6%, 29.9%]), followed by Help with Issues other than Primary Care (24.5% [22.9%, 26.1%]), Help with Adherence to ART (24.2% [22.4%, 26.1%]), and Help with Primary Care Appointments (23.1% [21.8%, 24.3%]). The range of importances for clients (17.8% - 32.0%, 14.2pp) was again larger than for providers (23.1% - 28.2%, 5.1pp). Help With Primary Care Appointments again had the biggest difference in relative importance between clients and providers (5.3pp lower among clients [−7pp, −3.5pp]).

### Part-worth Utilities

Figure 2 compares the part-worth utilities for clients and providers. Panel A shows the utilities under the scenario in which levels in the Where Program Visits Happen attribute were combined by location. Comparisons of the utilities between providers and clients were all statistically significant, with several differences in both the magnitude and direction of preference between populations. Within the Help with Adherence to ART attribute, clients preferred *Medication reminders via phone or text* (31.9 [25.4, 38.3]), followed by *Adherence assessment* (10.8 [4.9, 16.7]); they had a negative preference for *DOT* (−42.6 [−51, −34.3]). In contrast, *DOT* was the most preferred level for providers within the Help with Adherence to ART attribute (28 [20.8, 35.2]), with lower preferences for *Medication reminders via phone or text* (−5.3 [−11.3, 0.7]) and *Adherence assessment* (−22.7 [−30.3, −15]). Within the Help with Primary Care Appointments attribute, clients preferred *Appointment reminders and transportation* (12.5 [7.6, 17.5]), followed by *Appointment reminders* (0.9 [−3.9, 5.7]), with the lowest preference being for *Appointment reminders and accompaniment* (−13.4 [−18.5, −8.3]). However, *Appointment reminders and accompaniment* was the most preferred level for providers (21.4 [15.4, 27.4]), followed by *Appointment reminders and transportation* (20.3 [15.2, 25.4]), and *Appointment reminders* (−41.7 [−47.9, −35.5]). For the Help with Issues other than Primary Care attribute, *Housing and food* was the clients’ most preferred level (19.0 [12.4, 25.7]), followed by *Specialty medical care* (6.2 [0.6, 11.8]), *Mental health and well-being* (−12.4 [−17.8, −7]), and *Insurance, benefits, paperwork* (−12.8 [−17.8, −7.8]). In comparison, providers’ most preferred level was *Specialty medical care* (29 [23, 34.6]), followed by *Mental health and well-being* (18.8 [13.7, 23.9]), *Insurance, benefits, paperwork* (0.2 [−5.3, 5.6]), and *Housing and food* (−48 [−53, −43]). Finally, for Where Program Visits Happen, clients preferred *Meet via phone or video chat* (19.3 [12.9, 25.8]), followed by *Meet at home* (1.2 [−5.6, 8.1]), and *Meet at program location* (−20.6 [−28.2, −13]). Providers had the highest preference for *Meet at home* (19.4 [13.2, 25.5]), followed by *Meet at program location* (5.3 [−1, 11.6]), and the lowest preference for *Meet via phone or video chat* (−24.7 [−30.3, −19.1]). *DOT* had the largest utility gap between clients and providers (70.6 lower among clients [−81.6, −59.6]), followed by *Housing and food* (67.0 higher among clients [58.7, 75.3]), and *Meet via phone or video chat* (44.0 higher among clients ([35.5, 52.5]).

**Figure 2.**
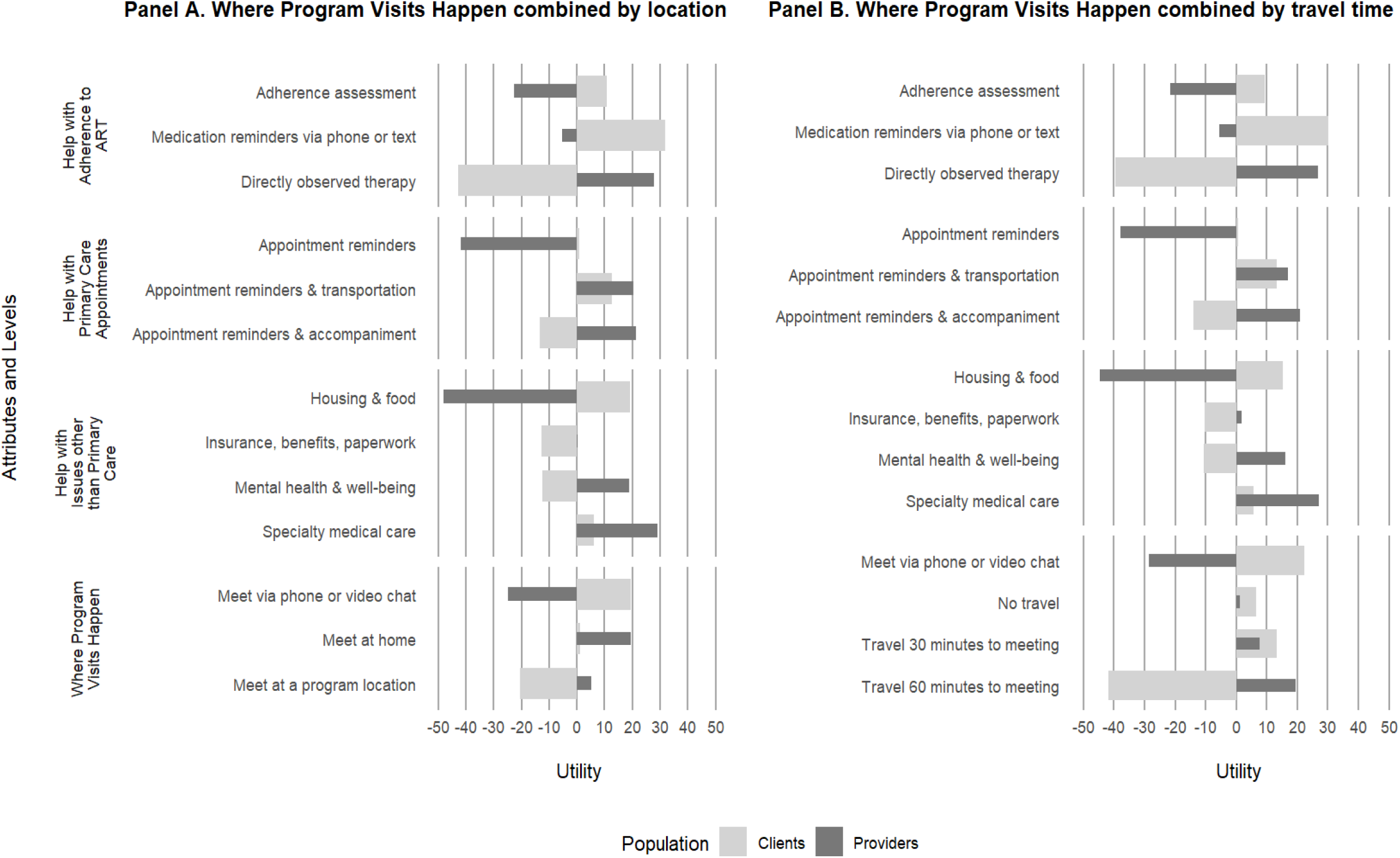
Part-worth utilities by analysis and population.

The patterns of preference and utility estimates in Panel B are similar to those in Panel A, with the exception of the Where Program Visits Happen attribute, here mapped by travel time. *Meet via phone or video chat* remained the most preferred level for clients (22.2 [15.2, 29.1]), but the next most preferred level was *Travel 30 minutes to meeting* (13.2 [7.8, 18.6]), then *No travel* (6.6 [−0.7, 13.9]) and lastly *Travel 60 minutes to meeting* (−41.9 [−49.7, −34.2]). For providers, *Travel 60 minutes to meeting* was the most preferred level (19.4 [10.1, 28.8]), *Travel 30 minutes to meeting* was the next most preferred level (7.8 [2.3, 13.3]), followed by *No travel* (1.3 [−5.4, 7.9]), and *Meet via phone or video chat* (−28.5 [−34.1, −22.9]). *DOT* again had the largest utility gap between clients and providers (66.4 lower among clients [−77.2, −55.5]), followed by *Travel 60 minutes to meeting* (61.4 lower among clients [−73.4, −49.3]) and *Housing and food* (60.0 higher among clients [52.9, 67.1]).

## Discussion

To our knowledge, this is the first study that has used a DCE to assess service provider-client agreement on non-clinical ART adherence support services. Other studies have largely focused on attributes of ART rather than ART adherence supports, and/or patient-physician comparisons rather than client-medical case management provider comparisons [20–22]. A recent survey of preferences for ART maintenance services among providers (including clinicians) and HIV-positive clients of healthcare facilities in Thailand used a standard multi-select survey question format [23]. These authors measured preferences for location, provider type, and the frequency of ART refills, viral load testing, STI testing, and psychosocial support, finding broad agreement for the most frequently selected category of each of the measures. This high level of agreement may derive from the type of survey question and the fact that respondents could pick all that apply; the focus on evaluation of preferences for clinical services; and/or cultural context. Another study focused on the degree of agreement between physicians and patients for characteristics of ART (for example, side effects and dosing flexibility) [20]. Notably, these authors used a DCE to assess patient preferences for these characteristics and to assess what physicians thought patient preferences were (rather than physicians’ own preferences), and they also found a high degree of agreement for the ART attributes included in the survey. Our study contributes to the literature by focusing on CCP providers and recipients and evaluating the concordance of their self-reported preferences for features of non-clinical adherence support services. Preference discordance could impact client satisfaction with services, retention in care, and ART adherence [12,13,24–27]. However, the present study only examined aggregate preferences and did not evaluate whether individual clients’ preferences were being met.

In the present study, client and provider preferences for CCP service intensity clearly diverged: clients tended to prefer lower-intensity services such as medication reminders via phone or text and virtual visits, whereas providers endorsed higher-intensity services such as home visits and accompanying clients to primary care appointments. While the magnitude of the preferences changed depending on the focus of the analysis, the direction of the preferences remained consistent across analyses within each group of respondents. Notably, in the location-based analysis, clients’ preference for program-based visits was strongly negative. In contrast, clients had a positive utility for 30 minutes of travel in the travel-time-based analysis and a strong negative utility for 60 minutes of travel, suggesting that it wasn’t the program location that influenced clients, but travel time.

There are several potential reasons for the discrepancies identified between client and provider preferences. Taking the survey, participating providers likely considered the range of clients’ needs across their caseloads and may have been enacting a belief that higher-intensity services are more likely to help their most vulnerable clients achieve ART adherence. Home visits, in particular, provide valuable information to staff about clients’ living conditions and how those may promote or inhibit adherence to treatment. In contrast, clients who participated in the DCE considered only their own needs. Participating clients had been enrolled in the CCP longer than their non-participating peers [3], and may not have perceived a need for such support for themselves, or may have felt that higher-intensity services such as DOT or home visits were intrusive or inconvenient. Providers may also have been motivated to endorse features of the CCP that they perceived as clearly distinguishing the CCP from other forms of medical case management.

Regardless of the reasons for discordant client and provider preferences, these results highlight the importance of engaging clients as partners in decisions about supportive services to make sure they are aligned with client values. It is also important to note that this analysis compares aggregated preferences across populations, and does not compare preferences within subgroups across populations. In our earlier work, latent class analyses identified subsets of clients who had higher preferences for higher-intensity services and providers who had higher preferences for lower-intensity services [3,5]. Our sample sizes required that these segmented analyses be exploratory in nature, which is why we did not explicitly consider heterogeneity in the present comparisons. However, those results alongside the present analysis support the 2018 revisions to the CCP, which allow for differentiated service delivery, increasingly recognized as central to improving client retention and adherence [28,29]

We recognize the following limitations of our study. The prompts used in the client and provider DCEs were the same: “Imagine that you had to choose between two programs with the features below. Select the one that you would prefer.” For clients, this prompt straightforwardly could be interpreted as pertaining to their preferences for themselves. For providers, it may have been unclear whether they were meant to make choices based on what program they would prefer to deliver, or what program they think would most benefit their clients (ambiguity that has been observed in concordance analyses in the context of other conditions as well [12]). However, as discussed in our previous analysis of provider preferences, we interpret the findings from the provider DCE as endorsement of the higher-intensity features of the CCP regardless of which perspective was motivating providers’ choices [12]. Future studies should carefully calibrate the instructions to each audience to make sure the desired framing is clear for each type of respondent.

In addition, the preferences of participating NYC CCP clients and providers are not expected to generalize to other jurisdictions. Nevertheless, we believe our results support differentiated service delivery, which has cross-cutting relevance, regardless of regional or other (e.g., urban-rural) differences in specific population needs or characteristics.

Lastly, the extent to which stated preferences, as measured via surveys such as DCEs, agree with revealed preferences, as indicated by actual choices and behaviors, is an area of ongoing inquiry [30–32]. Future work analyzing NYC Health Department records could add to the literature on this topic by measuring concordance between clients’ stated preferences and actual service utilization, and identifying predictors of agreement or divergence between stated and revealed preferences.

## Data Availability

Data is not publicly available.

## Acknowledgements

We gratefully acknowledge the contributions of Abigail Baim-Lance, Sarah Kozlowski, Tye Seabrook, Scarlett Macias, Graham Harriman, site coordinators, and the client and provider participants.

## Supplemental materials

**Supplemental Figure 1.**
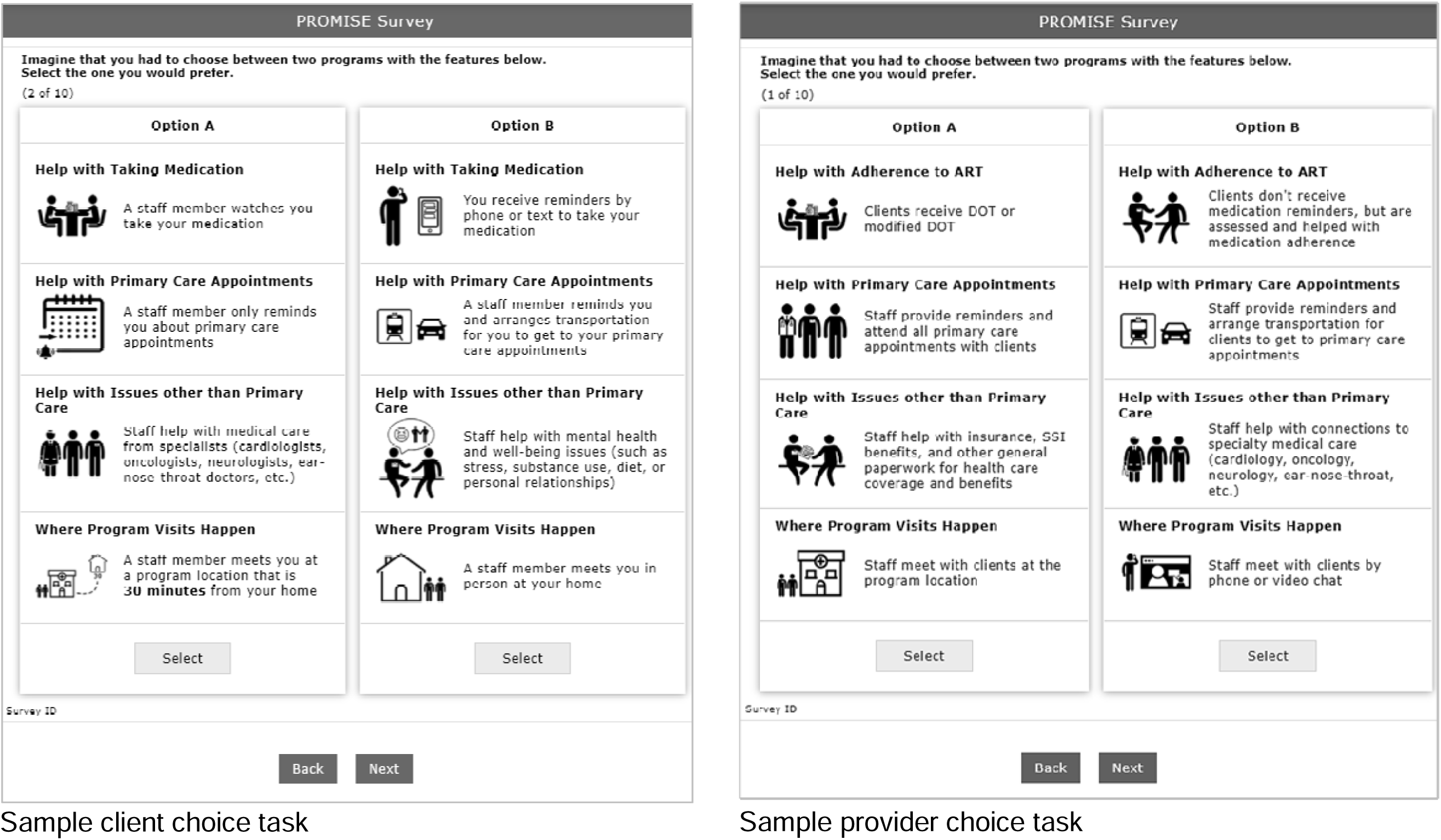

**Supplemental Table 1A.**
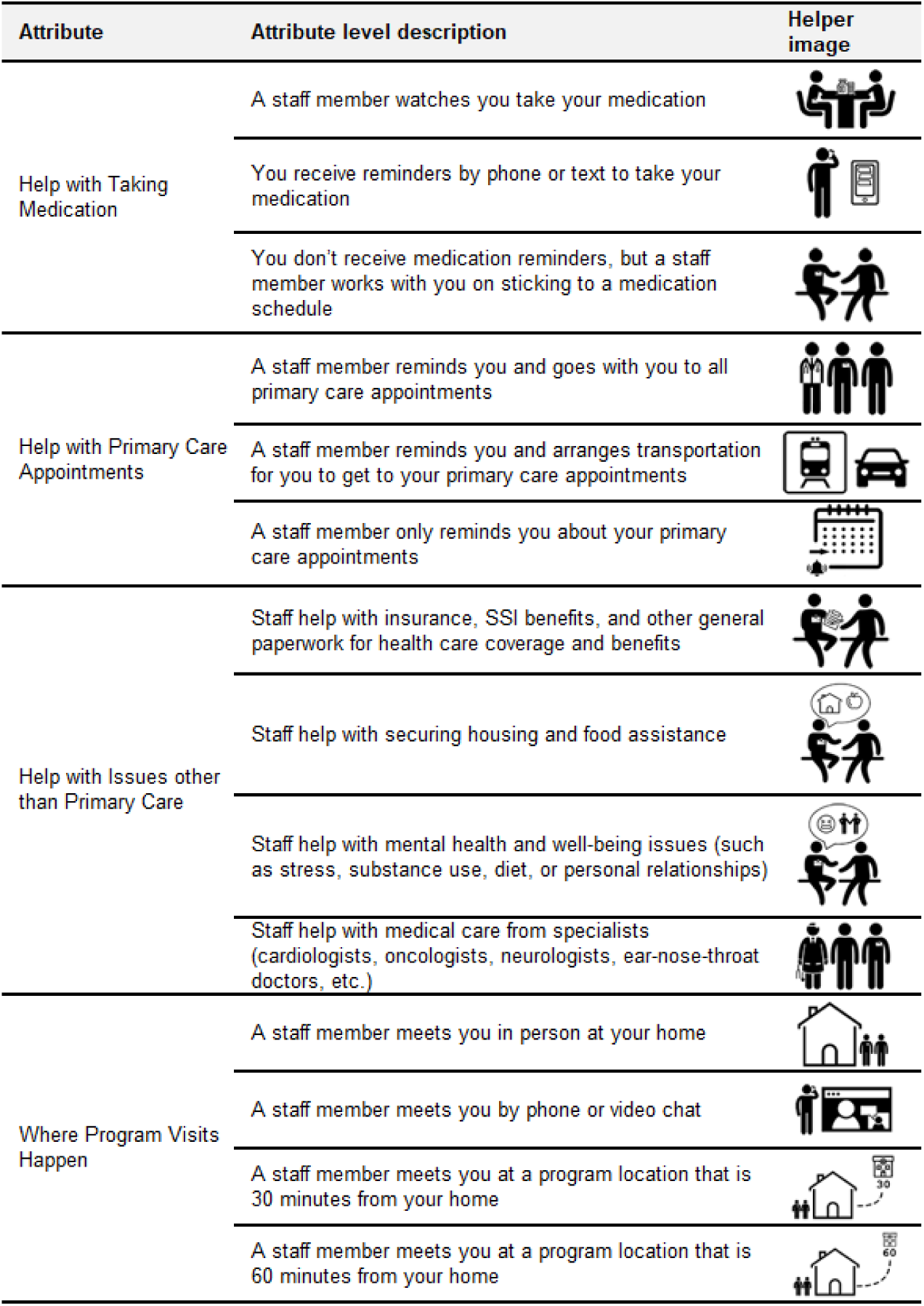
Attributes and levels of a discrete choice experiment investigating client preferences for HIV care coordination services in New York City.

| Attribute | Attribute level description | Helper image |
| --- | --- | --- |
| Help with Taking Medication | A staff member watches you take your medication |  |
|  | You receive reminders by phone or text to take your medication |  |
|  | You don't receive medication reminders, but a staff member works with you on sticking to a medication schedule |  |
| Help with Primary Care Appointments | A staff member reminds you and goes with you to all primary care appointments |  |
|  | A staff member reminds you and arranges transportation for you to get to your primary care appointments |  |
|  | A staff member only reminds you about your primary care appointments |  |
| Help with Issues other than Primary Care | Staff help with insurance, SSI benefits, and other general paperwork for health care coverage and benefits |  |
|  | Staff help with securing housing and food assistance |  |
|  | Staff help with mental health and well-being issues (such as stress, substance use, diet, or personal relationships) |  |
|  | Staff help with medical care from specialists (cardiologists, oncologists, neurologists, ear-nose-throat doctors, etc.) |  |
| Where Program Visits Happen | A staff member meets you in person at your home |  |
|  | A staff member meets you by phone or video chat |  |
|  | A staff member meets you at a program location that is 30 minutes from your home |  |
|  | A staff member meets you at a program location that is 60 minutes from your home |  |

**Supplemental Table 1B.**
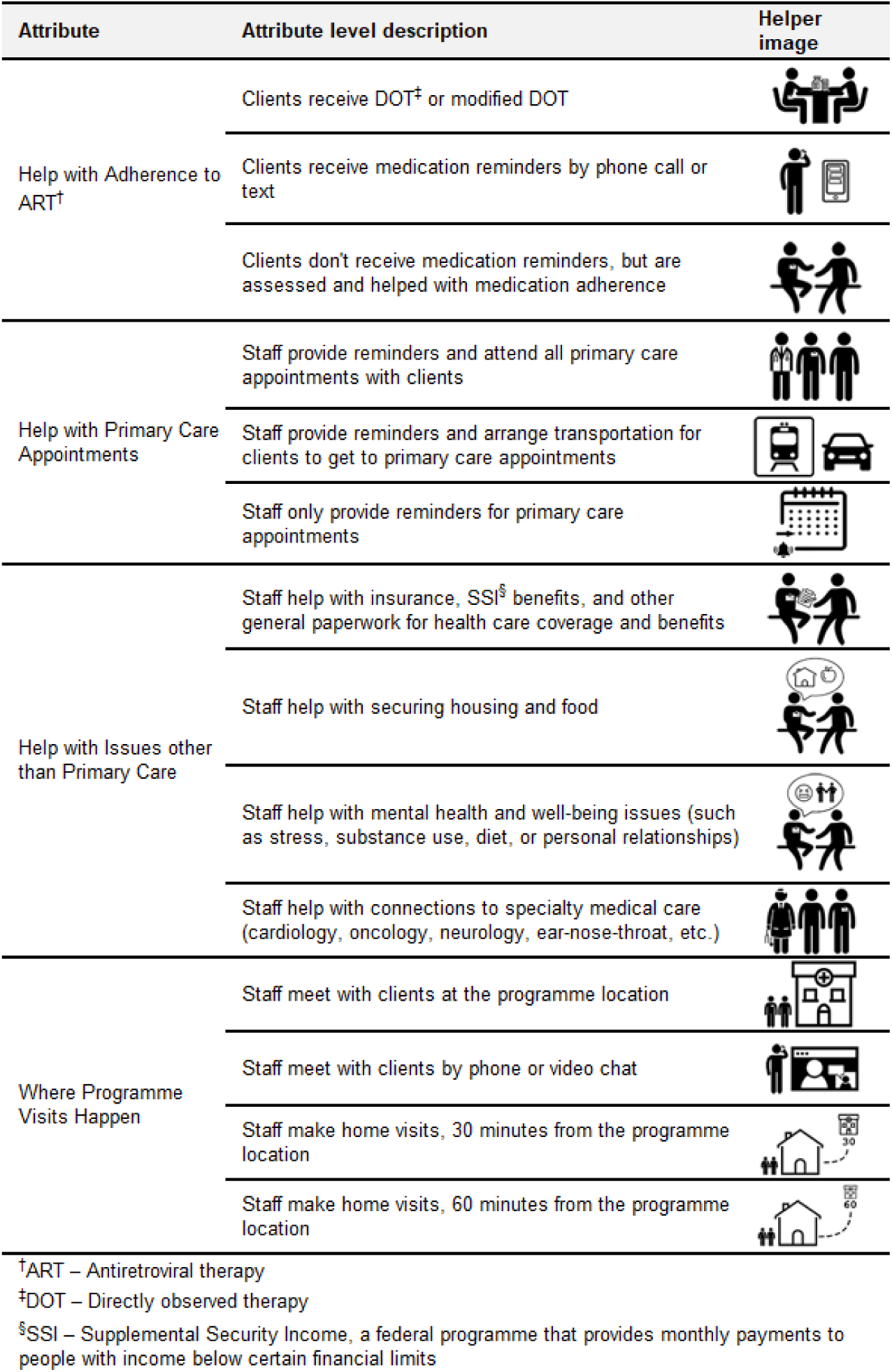
Attributes and levels of a discrete choice experiment investigating provider preferences for HIV care coordination services in New York City.

| Attribute | Attribute level description | Helper image |
| --- | --- | --- |
| Help with Adherence to ART<sup>†</sup> | Clients receive DOT<sup>‡</sup> or modified DOT |  |
|  | Clients receive medication reminders by phone call or text |  |
|  | Clients don't receive medication reminders, but are assessed and helped with medication adherence |  |
| Help with Primary Care Appointments | Staff provide reminders and attend all primary care appointments with clients |  |
|  | Staff provide reminders and arrange transportation for clients to get to primary care appointments |  |
|  | Staff only provide reminders for primary care appointments |  |
| Help with Issues other than Primary Care | Staff help with insurance, SSI<sup>§</sup> benefits, and other general paperwork for health care coverage and benefits |  |
|  | Staff help with securing housing and food |  |
|  | Staff help with mental health and well-being issues (such as stress, substance use, diet, or personal relationships) |  |
|  | Staff help with connections to specialty medical care (cardiology, oncology, neurology, ear-nose-throat, etc.) |  |
| Where Programme Visits Happen | Staff meet with clients at the programme location |  |
|  | Staff meet with clients by phone or video chat |  |
|  | Staff make home visits, 30 minutes from the programme location |  |
|  | Staff make home visits, 60 minutes from the programme location |  |

<sup>†</sup>ART – Antiretroviral therapy
<sup>‡</sup>DOT – Directly observed therapy
<sup>§</sup>SSI – Supplemental Security Income, a federal programme that provides monthly payments to people with income below certain financial limits

### Sensitivity Analyses

Out of the 181 total client responses, 140 (77.3%) were completed before the beginning of the COVID-19 pandemic. In the location-based analysis that included pre-COVID responses only, preference among clients for *Meet via phone or video chat* was lower (4.8 [−2.1, 11.8] versus 19.3 [12.9, 25.8]) than in the analysis that included all responses, while preferences for *Meet at home* and *Meet at program location* were higher (11.4 [3.8, 18.9] versus 1.2 [−5.6, 8.1], and −16.2 [−25.3, −7] versus −20.6 [−28.2, −13], respectively). In the travel time analysis that included pre-COVID client responses only, preference among clients for *Meet via phone or video chat* was also lower (11.4 [3.4, 19.4] versus 22.2 [15.2, 29.1]), while preference for *No travel* and *Travel 30 minutes to meeting* were higher (14.1 [6.5, 21.7] versus 6.6 [−0.7, 13.9], and 18.0 [11.3, 24.8] versus 13.2 [7.8, 18.6], respectively) compared to the analysis with all client responses. Preference for *Travel 60 minutes to meeting* remained approximately the same (−43.5 [−52.6, −34.5] pre-COVID client responses only and −41.9 [−49.7, −34.2] all client responses) regardless of when the surveys were completed. Since all provider responses were received before the start of the COVID-19 pandemic, provider preferences did not change.

